# Quantitative mobility measures complement the MDS-UPDRS for characterization of Parkinson’s disease heterogeneity

**DOI:** 10.1101/2020.08.16.20175596

**Authors:** Emily J. Hill, C. Grant Mangleburg, Isabel Alfradique-Dunham, Brittany Ripperger, Amanda Stillwell, Hiba Saade, Sindhu Rao, Oluwafunmiso Fagbongbe, Rainer von Coelln, Arjun Tarakad, Christine Hunter, Robert J. Dawe, Joseph Jankovic, Lisa M. Shulman, Aron S. Buchman, Joshua M. Shulman

## Abstract

**Introduction:** Emerging technologies show promise for enhanced characterization of Parkinson’s Disease (PD) motor manifestations. We evaluated quantitative mobility measures from a wearable device compared to the conventional motor assessment, the Movement Disorders Society-Unified PD Rating Scale part III (motor MDS-UPDRS).

**Methods:** We evaluated 176 subjects with PD (mean age 65, 65% male, 66% H&Y stage 2) at the time of routine clinic visits using the motor MDS-UPDRS and a structured 10-minute motor protocol, which included a 32-ft walk, Timed Up and Go (TUG), and standing posture with eyes closed, while wearing a body-fixed sensor (DynaPort MT, McRoberts BV). Regression models examined 12 quantitative mobility measures for associations with (i) motor MDS-UPDRS, (ii) motor subtype (tremor dominant vs. postural instability/gait difficulty), (iii) Montreal Cognitive Assessment (MoCA), and (iv) physical functioning disability (PROMIS-29). All analyses included age, gender, and disease duration as covariates. Models iii-iv were secondarily adjusted for motor MDS-UPDRS.

**Results:** Quantitative mobility measures from gait, TUG transitions, turning, and posture were significantly associated with motor MDS-UPDRS (7 of 12 measures, p< 0.05) and subtype (6 of 12 measures, p< 0.05). Compared with motor MDS-UPDRS, several quantitative mobility measures accounted for ∼1.5-fold increased variance in either cognition or physical functioning disability. Among minimally-impaired subjects within the bottom quartile of motor MDS-UPDRS, including subjects with normal gait exam, the measures captured substantial residual motor heterogeneity.

**Conclusion:** Clinic-based quantitative mobility assessments using a wearable sensor captured features of motor performance beyond those obtained with the motor MDS-UPDRS and may offer enhanced characterization of disease heterogeneity.

## 1. Introduction

Parkinson’s disease (PD) is diagnosed clinically, based on the presence of its cardinal motor manifestations, including bradykinesia, rigidity, gait/postural impairment, and tremor. The gold standard clinical research assessment tool for PD motor impairment is the Movement Disorder Society Unified Parkinson Disease Rating Scale (MDS-UPDRS) part III, a semi-quantitative measure consisting of 18 items measured on an ordinal scale ranging from 0 (normal) to 4 (severe) [1]. Although this scale is commonly used to measure motor symptom severity, individual patients often present with distinct PD motor profiles and these phenotypes can shift over time, reflecting a remarkable degree of heterogeneity [2–4]. Based on the MDS-UPDRS, for example, tremor dominant (TD) and postural instability/gait difficulty (PIGD) motor subtypes of PD have been recognized [5]. Non-motor manifestations, such as cognitive impairment, also variably contribute to disease heterogeneity and overall disability. Compared with conventional PD motor assessments, technology-enabled, quantitative mobility measures offer the promise of more sensitive detection, monitoring, and discrimination of heterogeneous phenotypes [6–8]. Several studies have demonstrated that quantitative mobility measures can detect motor features that distinguish PD from controls, including differences in gait [9–12], truncal movements [13–16], and the performance of motor tasks, such as the Timed Up and Go test (TUG) [17,18]. New outcome measures from devices may thus complement conventional evaluations by quantifying heterogeneous PD motor impairments.

Among the many emerging options for technology-enhanced PD outcome measures, some are limited to use in specialized gait labs, whereas others are designed for longer-term, unsupervised monitoring during everyday living [6,7]. The ability of wearable devices to enhance the assessment of PD mobility during routine clinical encounters has been less extensively studied. In prior work, we have validated quantitative mobility measures from a body-fixed sensor (DynaPort MT^®^, McRoberts BV) using a short, standardized motor protocol in older adults examined in the community setting [19,20]. In this sample, largely excluding subjects with PD, baseline quantitative mobility measures successfully predicted the development of mild parkinsonian signs [21]. Here, we deploy the same device to record similar motor tasks during routine PD clinical evaluations in our academic movement disorders outpatient practice setting and compare sensor-derived mobility metrics with the conventional motor MDS-UPDRS assessment.

## 2. Methods

### 2.1 Subjects and Clinical Evaluations

This study was approved by the Baylor College of Medicine Institutional Review Board. Subjects with PD were recruited during routine follow-up visits at the Baylor Parkinson’s Disease Center and Movement Disorders Clinic (PDCMDC). Any ambulatory patient able to complete the full assessment protocol was eligible for inclusion in the study. All subjects received a PD diagnosis based on evaluation by a movement disorders specialist. After informed consent, subjects received the following assessments: motor MDS-UPDRS (part III), the 10-minute quantitative motor protocol (see below), Montreal Cognitive Assessment (MoCA) and the Patient-Reported Outcomes Measurement Information System-29 profile (PROMIS-29). Current age and age at symptom onset were collected from the medical chart and confirmed with interview. The PROMIS-29 physical function score (including self-assessment of abilities in walking, shopping, and household chores) was calculated based on normative data [22]. For cohort descriptive purposes, we applied a cutoff of 40 or lower on the transformed ratio (T-score) of the physical function score to define subjects as disabled. Motor subtype (TD, PIGD, or indeterminate) was calculated from MDS-UPDRS subscores, using the established formula [23]. Subjects scoring in the indeterminate range (n = 16) were excluded from the analyses evaluating motor subtypes.

### 2.2 Gait and motor testing with a body-fixed sensor

The DynaPort MT is a small device (106.6 × 58 x 11.5mm, 55g) that is positioned at the low back in the midline affixed to a neoprene belt. Accelerometers and gyroscopes record data at a sampling frequency of 100 Hz, including for 3 acceleration axes (vertical, mediolateral, anteroposterior) and 3 angular velocity axes [yaw (rotation around the vertical axis); pitch (rotation around the mediolateral axis) and roll(rotation around the anteroposterior axis)]. As previously described [19], subjects performed a 10-minute motor protocol while wearing the DynaPortMT device, consisting of three tasks: (i) 32-foot walk (subjects walk a distance of eight feet four times without stopping, including turns); (ii) the TUG [subjects stand from a chair, walk eight feet at a comfortable pace, turn (turn 1), walk back to the chair, turn again (turn 2), and sit down, after one practice trial]; and (iii) standing posture with eyes closed for 20 seconds. This protocol was designed to be feasible to perform in an enclosed space. The device connects wirelessly to a laptop computer, enabling a research assistant to control data collection. Following the motor protocol, data is transferred from the device to the laptop and then uploaded to a central server for post-hoc segmentation of individual performances, quality control procedures, and extraction of measures using established algorithms within a MATLAB software package (MathWorks, Natick, MA). The derivation and validation of these scores were detailed in prior publications [18,24]. The 12 scores (referred to below as quantitative mobility measures) were derived from the 3 motor tasks as follows: (i) 4 measures from the 32-foot walk, (ii) 7 from TUG (3 from sitting to standing transition (S1), 2 from standing to sitting transition (S2), and 2 from turning), and (iii) 1 from standing posture with eyes closed. All measures were standardized with mean = 0 and standard deviation = 1. Outliers over 3 standard deviations from the mean for each calculated measure were excluded, and transformations for normality were applied as in our prior work [18,24].

### 2.3 Statistical analysis

All statistical analyses were performed using R software (http://www.R-project.org/). Due to variable missingness for the 12 mobility measures, total subject sample size for each analysis ranged from 149 to 176 (Table S1). Linear regression was first used to analyze relationships between the 12 individual quantitative mobility measures and (i) motor MDS-UPDRS, (ii) MoCA score, and (iii) PROMIS physical functioning disability. Logistic regression was similarly used to evaluate associations with (iv) PD motor subtype. All analyses were adjusted for age, sex, and disease duration as covariates. We next repeated analyses ii & iii including motor MDS-UPDRS as a covariate and examined whether the associations between quantitative mobility measures and each outcome remained significant. In secondary analyses, regression models were repeated including a covariate for subject-reported on/off dopaminergic medication clinical status (MDS-UPDRS item 3b). Joint regression models were also constructed to evaluate the variance in either cognition (MoCA) or physical functioning disability (PROMIS-29) explained by multiple, independently-associated mobility measures. The adjusted R-squared for a base model (including terms for age, sex, and disease duration) was compared to models including additional terms for the motor MDS-UPDRS with/without the mobility measures [20]. Lastly, we created violin plots to visualize the distribution of conventional motor measurements (motor MDS-UPDRS) compared to that from quantitative mobility measures. Measurement values were normalized to a range from 0 to 1 using the formula: (x-x_min_)/(x_max_-x_min_), where x is the measurement value being normalized, x_min_ is the minimum value amongst all measurements for that metric, and x_max_ is the maximum value amongst all measurements for that metric. Following normalization, violin and superimposed dot plots were generated using the “ggplot2” package in R, including a horizontal and vertical jitter.

## 3. Results

Following quality control filters, our study included 176 subjects with PD. Cohort clinical and demographic characteristics are summarized in Table 1, and descriptive statistics for the 12 quantitative mobility measures are displayed in the supplement (Table S1). The majority of subjects (91%) reported no or only mild motor fluctuations (0–25% off time per day).

**Table 1:**
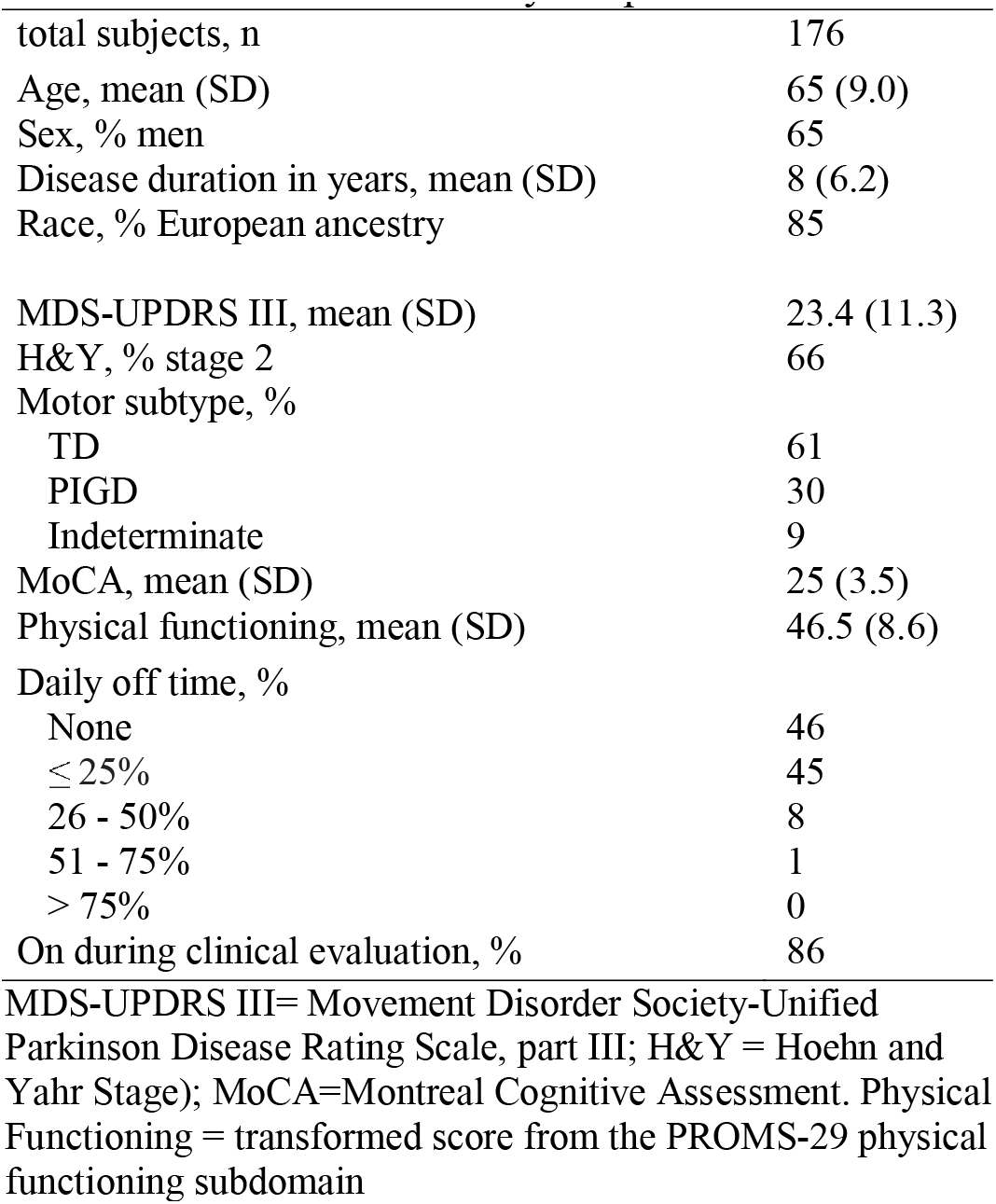
Clinical characteristics of study sample

We first examined associations between each of the DynaPort mobility measures and the motor MDS-UPDRS. Seven of 12 mobility measures were significantly associated with the motor MDS-UPDRS after adjustment for age, sex, and disease duration (Table 2). This includes measures from the 32-ft walk (speed, p< 0.001, cadence, p< 0.05, and stride variability, p< 0.01), the TUG sit to stand transition (S1 posterior, p< 0.01), the TUG stand to sit transition (S2 jerk, p< 0.01), turning (yaw, p< 0.0001), and standing posture with eyes closed (sway, p< 0.001) (Table 2). Conversely, the remaining measures were not significantly associated with the motor MDS-UPDRS. Six of the 12 mobility measures were also significantly associated with PD motor subtype (Table S2), including S1 range from the TUG and 4 other measures that were also associated with motor MDS-UPDRS. When compared with TD, subjects with the PIGD PD motor subtype were characterized by reduced speed and increased stride variability during the 32-ft walk task, reduced truncal movements during the TUG [S1 range, S1 posterior, and yaw], and increased sway during standing posture. Nearly all associations remained significant after adjustment for subject-reported on/off dopaminergic medication clinical status (Table S3).

**Table 2:**
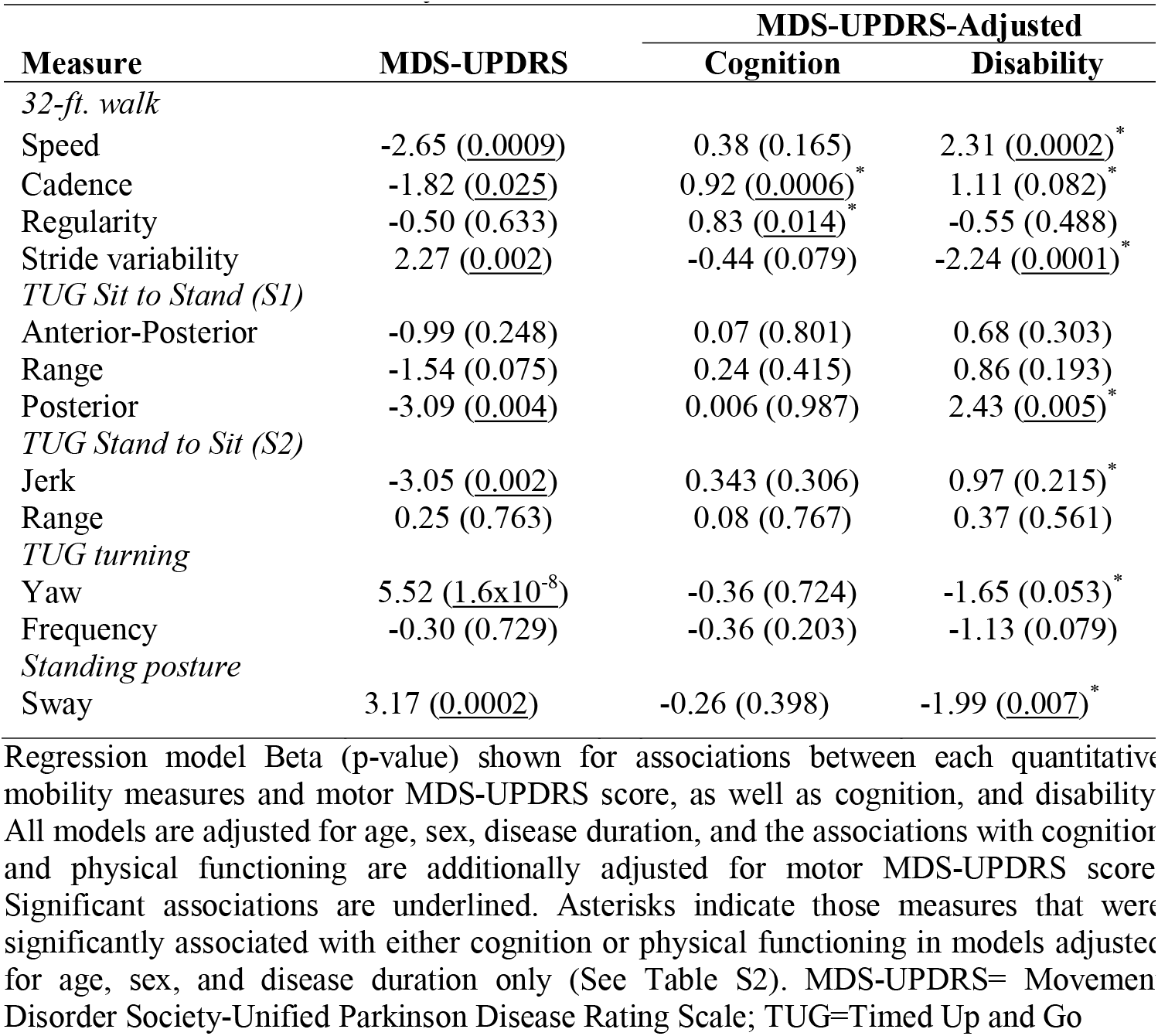
Associations between mobility measures and PD outcomes

PD progression is characterized by greater motor impairment over time; consequently, increasing motor MDS-UPDRS score is commonly related to similar worsening in cognitive impairment and disease-related disability [25]. Such correlated but independent, clinically-relevant PD outcome measures can serve as useful “anchors” for head-to-head comparison of the DynaPort quantitative mobility measures and conventional assessments using the motor MDS-UPDRS. Therefore, we next compared the quantitative mobility measures and the motor MDS-UPDRS for their associations with cognition and physical functioning disability. Mean MoCA score was 25 (SD = 3.5) in our sample, and 19.5% were classified as having significant disability (see Methods). As expected, similar to motor MDS-UPDRS, several quantitative mobility measures were also associated with cognition (2 measures) and/or physical functioning disability (7 measures) (Table S2). In order to determine if the mobility measures were independently associated with cognition and disability, we repeated these analyses including motor MDSUPDRS score as a covariate. Indeed, the associations between cognition and 2 measures from the 32-ft walk task (cadence and regularity) were each robust to adjustment for motor MDS-UPDRS score. Moreover, 4 out of 7 mobility measures remained significantly associated with disability following adjustment for motor MDS-UPDRS (Table 2).

Next, in order to quantify the information provided by the mobility measures that is not provided by motor MDS-UPDRS, we examined joint regression models for cognition and disability (Table 3). Our base model, including the variables for age, sex, and disease duration, accounted for 7.5% and 10% of the variance in cognition and physical functioning disability, respectively. Adding the motor MDS-UPDRS score to the model captured 5.2% more variance in cognition. Adding the 2 quantitative mobility measures that were associated with cognition independent of motor MDS-UPDRS accounted for an additional 6.7% variance, representing a 1.5-fold increase. Similarly, in the joint model for disability, the 4 independently-associated, quantitative mobility measures accounted for an additional 13% variance in the disability outcome—a 1.7-fold increase over adding motor MDS-UPDRS alone. Our findings were overall consistent following adjustment for patient-reported on/off dopaminergic medication status (Table S4).

**Table 3:** Joint Regression Models for Cognition and Physical Functioning Disability

| Variable | Cognition |  |  | Disability |  |  |
| --- | --- | --- | --- | --- | --- | --- |
|  | Model A | Model B | Model C | Model A | Model B | Model C |
| Age | -0.117 (7.15x10<sup>-5</sup>) | -0.085 (0.005) | -0.064 (0.048) | -0.138 (0.048) | -0.042 (0.55) | 0.024 (0.72) |
| Sex | -0.237 (0.663) | 0.203 (0.709) | 0.376 (0.508) | 1.681 (0.197) | 2.984 (0.02) | 0.564 (0.68) |
| Disease Duration | 0.017 (0.687) | 0.047 (0.264) | 0.039 (0.376) | -0.407 (8.03x10<sup>-5</sup>) | -0.319 (0.001) | -0.354 (0.005) |
| MDS-UPDRS III |  | -0.083 (0.001) | -0.074 (0.005) |  | -0.247 (4.2 x10<sup>-5</sup>) | -0.068 (0.29) |
| Speed |  |  |  |  |  | 1.761 (0.007) |
| Cadence |  |  | 0.786 (0.007) |  |  |  |
| Step Regularity |  |  | 0.78 (0.02) |  |  |  |
| Stride Variability |  |  |  |  |  | -1.148 (0.07) |
| Posterior |  |  |  |  |  | 1.857 (0.04) |
| Sway |  |  |  |  |  | -1.914 (0.007) |
| Adjusted R<sup>2</sup> | 0.075 | 0.127 | 0.194 | 0.100 | 0.180 | 0.310 |
| Fold increase |  | 1.7 | 1.5 |  | 1.8 | 1.7 |

Beta (p-value) shown from regression models examining cognition or physical functioning disability as an outcome. The base model (Model A) includes age, sex, and disease duration as covariates. In Model B, motor MDS-UPDRS III is added as a covariate. Model C includes all prior covariates as well as all quantitative mobility measures that were found to have individual independent associations with the outcome. Each model's adjusted R-squared is displayed at the bottom of the column and the amount of increase in the R-squared in Models B and C are displayed below. MDS-UPDRS III = Movement Disorders Society-Unified Parkinson Disease Rating Scale, part III; S1= Sit to stand transition in the Timed up and Go.

These results strongly suggest that quantitative mobility measures from a wearable device capture information about PD motor impairment that is distinct and complementary to the motor MDS-UPDRS. In order to better understand the relationship between these outcomes, we plotted the normalized distribution of data points for both measures, allowing for direct visual comparison. As displayed in Figure 1A, subjects falling within the upper (red) and lower (green) quartiles of the motor MDS-UPDRS, are observed to redistribute among all 4 quartiles of the normalized distribution for multiple mobility measures. For example, among the individuals clustered tightly in the bottom quartile of motor MDSUPDRS, some were found to have correspondingly high stride variability measures, whereas others fall in the middle or low range of values. Thus, stride variability and several other quantitative mobility measures can stratify PD subjects based on differences in motor performance that are not appreciable based on total motor MDS-UPDRS score alone. We next compared the score from the gait item of the motor MDS-UPDRS (item 3.10) to the normalized distribution of the quantitative mobility measures derived from the 32-ft walk task (Figure 1B). The MDS-UPDRS gait item is scored on an ordinal scale (0–4). In our sample, 89% of subjects were characterized by mild or absent gait impairment (score of 0–1) based on the motor MDS-UPDRS gait item. However, quantitative gait assessment using the device revealed substantial heterogeneity in gait performance among these same subjects. In fact, among the 75 subjects without detectable gait impairment on the MDS-UPDRS (gait score = 0, black in Figure 1B), quantitative measures of speed (mean = –0.12, SD = 0.995), cadence (mean = –0.09, SD = –0.98), regularity (mean = 0.02, SD = 0.81), and stride variability (mean = 0.06, SD = 1.05) each captured substantial variation, thereby providing a more granular assessment of gait.

**Figure 1.**
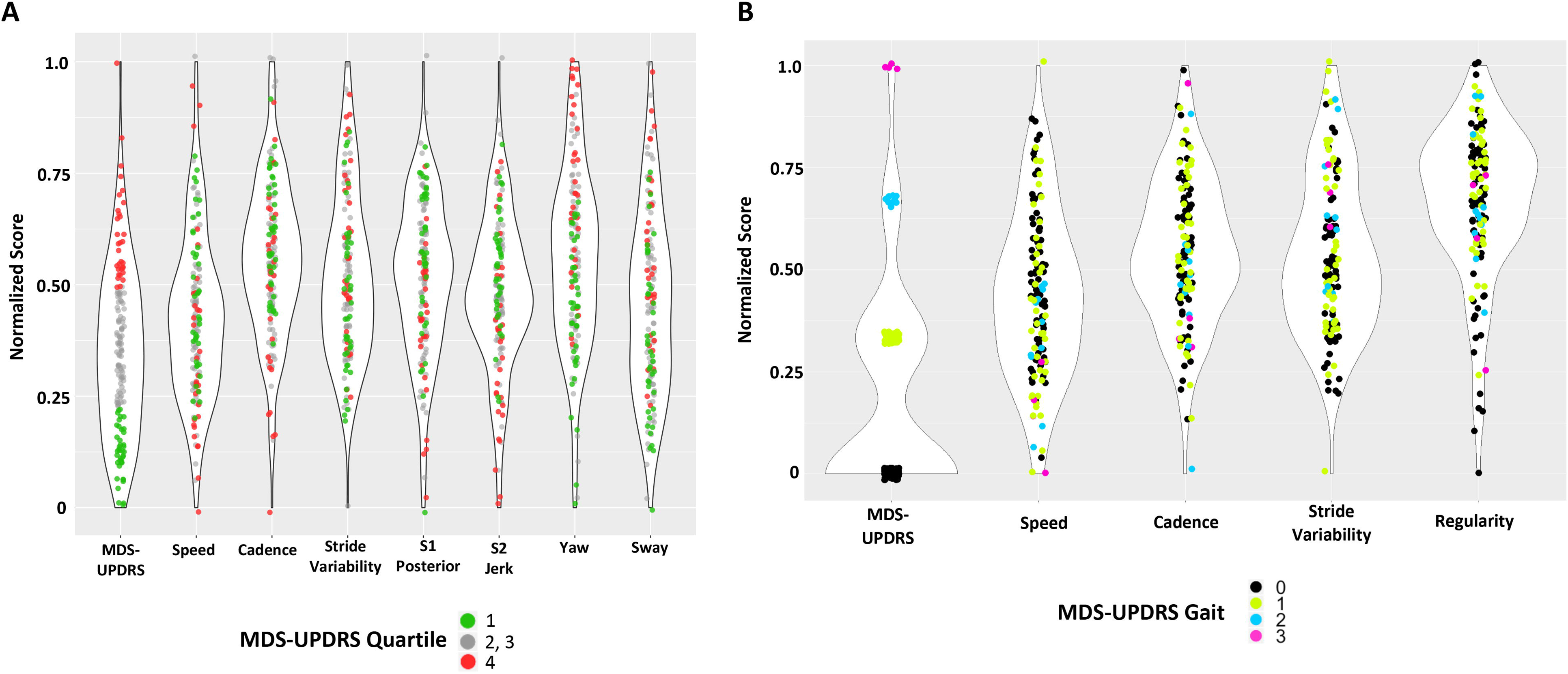
Violin plots of motor MDS-UPDRS versus quantitative mobility measures. The distribution of subject motor MDS-UPDRS scores is compared with that for quantitative mobility measures. Low scores are normalized to 0 and high scores are normalized to 1 to facilitate comparison. The width of the violin represents the number of data points at that value. (A) Total motor MDS-UPDRS score is compared with the 7 associated quantitative mobility measures (Table 2). Subjects falling within the upper and lower quartile of MDS-UPDRS are displayed in red and green, respectively. (B) The MDSUPDRS gait item is compared with the quantitative mobility measures from the 32-ft walk task. Subjects are displayed with colors based on the MDS-UPDRS gait item score: 0, black; 1, green; 2, blue; 3, magenta.

## 4. Discussion

The motor MDS-UPDRS is the gold standard battery for monitoring PD motor progression and response to therapies. Emerging quantitative motor assessments require evaluation of how these new outcome measures can complement conventional assessments, such as the MDS-UPDRS. During routine office visits, we obtained quantitative mobility measures in 176 subjects with PD, based on completion of several motor tasks while wearing the DynaPort device. Several of the quantitative mobility measures, including measures from the 32-ft walk (speed, cadence, and stride variability), the TUG (S1 posterior, S2 jerk, yaw), and standing posture (sway), were significantly associated with motor MDS-UPDRS. This result is consistent with other studies deploying devices for PD monitoring [26], and it is not surprising since the MDS-UPDRS includes semi-quantitative assessments of PD motor features that overlap with our protocol, including examination of gait (item 3.10–3.11) and arising from a chair (3.9). By contrast, several other quantitative mobility measures, were not significantly associated with motor MDS-UPDRS, suggesting they may capture independent features of PD motor impairment. In fact, several features of motor impairment in PD, such as turning, are not directly scored by the MDS-UPDRS. Of the 2 measures derived from turning in our analyses, only one (yaw) was associated with motor MDS-UPDRS score, whereas frequency was not significantly associated. The DynaPort device records data at 100Hz and records multiple independent measures for selected tasks, thus providing a more granular assessment of mobility. For example, whereas the motor MDS-UPDRS integrates multiple dimensions of gait into a single categorical score, our analyses considered 4 continuous measures from the 32-ft walk (speed, cadence, regularity, and stride variability). Indeed, following adjustment for motor MDS-UPDRS, we found that several DynaPort mobility measures remained significantly associated with cognition and/or disability, and these measures explained additional variance for these outcomes in joint regression models. Overall, our results highlight how quantitative mobility measures from a wearable device can complement conventional assessments, characterizing relevant features of PD motor impairment that are not adequately captured using the motor MDS-UPDRS.

Among subjects in our cohort with little or no gait impairment based on conventional rating scale evaluations, we found that several quantitative mobility measures revealed substantial variation in performance. Given that PD is preceded by a prolonged clinical prodromal phase, improved mobility biomarkers are urgently needed for sensitive and robust detection of the earliest PD-related motor manifestations [27]. In one study that also used the DynaPort, stride time variability during a 20-meter gait task successfully identified carriers of a dominant *LRRK2* (G2019S) allele who were not diagnosed with PD based on neurologic evaluation including the UPDRS [28]. In another “at-risk” cohort, gait mat evaluations revealed differences in step cadence and swing time among subjects with REM sleep behavior disorder, similarly with little or no impairment based on motor UPDRS scores [29]. Further, in a longitudinal study of subjects without clinically recognized disease, step time variability derived from baseline DynaPort gait evaluations predicted conversion to PD within 5 years [30]. Lastly, using an identical assessment protocol as in our present work, we found that baseline difference in quantitative mobility measures can also predict the development of mild parkinsonian signs, based on a modified motor UPDRS, in older adults without PD [21]. In sum, our results add to other published work showing how quantitative assessments using devices are more sensitive than the motor MDS-UPDRS for detecting gait alterations, and may serve as a useful biomarker for early (or prodromal) PD.

Besides its subtle onset, PD is characterized by substantial heterogeneity in the specific combinations and severity of motor, as well as non-motor features, with implications for prognosis and disease-related disability [2]. Optimal mobility biomarkers must therefore not only achieve high sensitivity for early detection, but also permit differentiation of distinct inter-individual patterns of motor impairment [31]. Six out of 12 quantitative mobility measures were associated with PD motor subtype characterized by TD versus PIGD predominant features. Moreover, selected mobility measures, including step cadence and regularity during the gait task, strongly associated with PD cognitive impairment, an important source of non-motor heterogeneity. Importantly, by considering subjects in the top or bottom quartile based on the motor MDS-UPDRS, we found that multiple DynaPort mobility measures detected substantial residual variation. In other words, these measures reveal “latent heterogeneity” among PD subjects—variation in motor performance among individuals that appear similarly affected based on total motor MDS-UPDRS score. In the future, motor assessments using wearable or other devices may thus enhance patient stratification based on PD motor subtypes [6,32].

Strengths of our study include a large, clinic-based PD sample with systematic motor evaluations using both MDS-UPDRS and wearable devices. Moreover, our previously-validated, short motor protocol [19,21] was deployed during routine clinic visits without requirement of a specialized gait laboratory. We also acknowledge a number of important limitations. Since our study population included predominantly early PD with mild overall impairment, it will be important in future studies to recruit a larger sample including the full spectrum of disease and to perform longitudinal evaluations to directly monitor progression. Although adjustment for subject-reported dopaminergic medication status did not significantly affect our findings, it may be informative to perform on/off evaluations in selected subjects along with repeated assessments to better understand intraindividual variability of these measures, as motor symptoms in PD often fluctuate. Lastly, our reliance on a single device (DynaPort), motor protocol, and its derived measures potentially limits generalizability, and our analyses excluded direct assessment of certain relevant PD motor features, such as tremor or dyskinesia. Our findings nevertheless establish how wearable devices can complement the motor MDS-UPDRS and offer enhanced motor phenotyping for PD.

## Data Availability

All data referred to in the manuscript is included in the manuscript tables & figures and/or is available on request from the corresponding author.

## Acknowledgements

The authors would like to thank Caitlin Zasadil, MS at Rush University Medical Center for her assistance with data processing and analysis. EJH was supported by the Parkinson Study Group / Parkinson’s Foundation’s Mentored Clinical Research Award and the National Human Genome Research Institute Medical Genetics Research Fellowship (T32GM007526–41). JMS was supported by Huffington Foundation and a Career Award for Medical Scientists from the Burroughs Welcome Fund. LMS was supported by the NIH and the Rosalyn Newman Foundation. RJD was supported by the National Institute on Aging Mentored Quantitative Research Development Award (K25AG61254). AB received support from NIH (R01AG056352, R01AG017917, RF1AG022018). JJ received research/training funding from AbbVie Inc, CHDI Foundation, Dystonia Coalition, Hoffmann-La Roche Ltd, Michael J Fox Foundation for Parkinson Research, National Institutes of Health, Parkinson’s Foundation, Parkinson Study Group, Roche, and Teva Pharmaceutical Industries Ltd.

## References

[1] C.G. Goetz, G.T. Stebbins, L.M. Blasucci, Differential progression of motor impairment in levodopa-treated Parkinson’s disease, Mov. Disord. (2000). https://doi.org/10.1002/1531-8257(200005)15:3<479::AID-MDS1009>3.0.CO;2-P.

[2] M.A. Thenganatt, J. Jankovic, Parkinson disease subtypes, JAMA Neurol. 71 (2014) 499–504. https://doi.org/10.1001/jamaneurol.2013.6233.

[3] R. Von Coelln, L.M. Shulman, Clinical subtypes and genetic heterogeneity: Of lumping and splitting in Parkinson disease, Curr. Opin. Neurol. (2016). https://doi.org/10.1097/WCO.0000000000000384.

[4] T. Simuni, C. Caspell-Garcia, C. Coffey, S. Lasch, C. Tanner, K. Marek, How stable are Parkinson’s disease subtypes in de novo patients: Analysis of the PPMI cohort?, Park. Relat. Disord. 28 (2016) 62–67. https://doi.org/10.1016/j.parkreldis.2016.04.027.2016.04.027.

[5] Jankovic J, McDermott M, Carter J, Gauthier S, Goetz C, Golbe L, Huber S, Koller W, Olanow C, Shoulson I, et al. Variable expression of Parkinson’s disease: a base-line analysis of the DATATOP cohort. The Parkinson Study Group. Neurology. 1990 Oct;40(10):1529–34. doi: 10.1212/wnl.40.10.1529.PMID:2215943

[6] A.J. Espay, J.M. Hausdorff, Á. Sánchez-Ferro, J. Klucken, A. Merola, P. Bonato, S.S. Paul, F.B. Horak, J.A. Vizcarra, T.A. Mestre, R. Reilmann, A. Nieuwboer, E.R. Dorsey, L. Rochester, B.R. Bloem, W. Maetzler, A roadmap for implementation of patient-centered digital outcome measures in Parkinson’s disease obtained using mobile health technologies, Mov. Disord. 34 (2019) 657–663. https://doi.org/10.1002/mds.27671.

[7] C.A. Artusi, M. Mishra, P. Latimer, J.A. Vizcarra, L. Lopiano, W. Maetzler, A. Merola, A.J. Espay, Integration of technology-based outcome measures in clinical trials of Parkinson and other neurodegenerative diseases, Park. Relat. Disord. 46 (2018) S53–S56. https://doi.org/10.1016/j.parkreldis.2017.07.022.

[8] A. Tarakad, Clinical Rating Scales and Quantitative Assessments of Movement Disorders, Neurol. Clin. 38 (2020) 231–254. https://doi.org/10.1016/j.ncl.2019.12.001.

[9] R. Baltadjieva, N. Giladi, L. Gruendlinger, C. Peretz, J.M. Hausdorff, Marked alterations in the gait timing and rhythmicity of patients with de novo Parkinson’s disease, Eur. J. Neurosci. 24 (2006) 1815–1820. https://doi.org/10.1111/j.1460-9568.2006.05033.x.

[10] J.M. Hausdorff, Gait dynamics in Parkinson’s disease: Common and distinct behavior among stride length, gait variability, and fractal-like scaling, Chaos. 19 (2009). https://doi.org/10.1063/1.3147408.

[11] F.B. Horak, M. Mancini, Objective biomarkers of balance and gait for Parkinson’s disease using body-worn sensors, Mov. Disord. 28 (2013) 1544–1551. https://doi.org/10.1002/mds.25684.

[12] J.C.M. Schlachetzki, J. Barth, F. Marxreiter, J. Gossler, Z. Kohl, S. Reinfelder, H. Gassner, K. Aminian, B.M. Eskofier, J. Winkler, J. Klucken, Wearable sensors objectively measure gait parameters in Parkinson’s disease, PLoS One. 12 (2017). https://doi.org/10.1371/journal.pone.0183989.

[13] M. Mancini, C. Zampieri, P. Carlson-Kuhta, L. Chiari, F.B. Horak, Anticipatory postural adjustments prior to step initiation are hypometric in untreated Parkinson’s disease: An accelerometer-based approach, Eur. J. Neurol. 16 (2009) 1028–1034. https://doi.org/10.1111/j.1468-1331.2009.02641.x.

[14] M. Mancini, P. Carlson-Kuhta, C. Zampieri, J.G. Nutt, L. Chiari, F.B. Horak, Postural sway as a marker of progression in Parkinson’s disease: A pilot longitudinal study, Gait Posture. 36 (2012) 471–476. https://doi.org/10.1016/j.gaitpost.2012.04.010.

[15] W. Maetzler, M. Mancini, I. Liepelt-Scarfone, K. Müller, C. Becker, R.C. van Lummel, E. Ainsworth, M. Hobert, J. Streffer, D. Berg, L. Chiari, Impaired trunk stability in individuals at high risk for Parkinson’s disease, PLoS One. 7 (2012). https://doi.org/10.1371/journal.pone.0032240.

[16] S.J. Ozinga, A.G. Machado, M. Miller Koop, A.B. Rosenfeldt, J.L. Alberts, Objective assessment of postural stability in Parkinson’s disease using mobile technology, Mov. Disord. (2015). https://doi.org/10.1002/mds.26214.

[17] C. Zampieri, A. Salarian, P. Carlson-Kuhta, K. Aminian, J.G. Nutt, F.B. Horak, The instrumented timed up and go test: Potential outcome measure for disease modifying therapies in Parkinson’s disease, J. Neurol. Neurosurg. Psychiatry. (2010). https://doi.org/10.1136/jnnp.2009.173740.

[18] A. Weiss, T. Herman, M. Plotnik, M. Brozgol, I. Maidan, N. Giladi, T. Gurevich, J.M. Hausdorff, Can an accelerometer enhance the utility of the Timed Up & Go Test when evaluating patients with Parkinson’s disease?, Med. Eng. Phys. (2010). https://doi.org/10.1016/j.medengphy.2009.10.015.2009.10.015.

[19] A.S. Buchman, S.E. Leurgans, A. Weiss, V. VanderHorst, A. Mirelman, R. Dawe, L.L. Barnes, R.S. Wilson, J.M. Hausdorff, D.A. Bennett, Associations between quantitative mobility measures derived from components of conventional mobility testing and parkinsonian gait in older adults, PLoS One. (2014). https://doi.org/10.1371/journal.pone.0086262.

[20] R.J. Dawe, S.E. Leurgans, J. Yang, J.M. Bennett, J.M. Hausdorff, A.S. Lim, C. Gaiteri, D.A. Bennett, A.S. Buchman, Association between Quantitative Gait and Balance Measures and Total Daily Physical Activity in Community-Dwelling Older Adults, Journals Gerontol. – Ser. A Biol. Sci. Med. Sci. 73 (2018) 636–642. https://doi.org/10.1093/gerona/glx167.

[21] R. von Coelln, R.J. Dawe, S.E. Leurgans, T.A. Curran, T. Truty, L. Yu, L.L. Barnes, J.M. Shulman, L.M. Shulman, D.A. Bennett, J.M. Hausdorff, A.S. Buchman, Quantitative mobility metrics from a wearable sensor predict incident parkinsonism in older adults, Park. Relat. Disord. 65 (2019) 190–196. https://doi.org/10.1016/j.parkreldis.2019.06.012.2019.06.012.

[22] S.E. Jensen, D. Cella, P.A. Pilkonis, B.B. Reeve, S. Czajkowski, A.A. Stone, D. Amtmann, D.A. Dewalt, K.P. Weinfurt, J.F. Fries, B.D. Schalet, K.F. Cook, J.L. Beaumont, PROMIS measures of pain, fatigue, negative affect, physical function, and social function demonstrated clinical validity across a range of chronic conditions, J. Clin. Epidemiol. (2016). https://doi.org/10.1016/j.jclinepi.2015.08.038.2015.08.038.

[23] G.T. Stebbins, C.G. Goetz, D.J. Burn, J. Jankovic, T.K. Khoo, B.C. Tilley, How to identify tremor dominant and postural instability/gait difficulty groups with the movement disorder society unified Parkinson’s disease rating scale: Comparison with the unified Parkinson’s disease rating scale, Mov. Disord. 28 (2013) 668–670. https://doi.org/10.1002/mds.25383.

[24] A. Weiss, T. Herman, M. Plotnik, M. Brozgol, N. Giladi, J.M. Hausdorff, An instrumented timed up and go: The added value of an accelerometer for identifying fall risk in idiopathic fallers, Physiol. Meas. (2011). https://doi.org/10.1088/0967-3334/32/12/009.

[25] J. Jankovic, A.S. Kapadia, Functional decline in Parkinson disease, Arch. Neurol. 58 (2001) 1611–1615. https://doi.org/10.1001/archneur.58.10.1611.

[26] B. Boroojerdi, R. Ghaffari, N. Mahadevan, M. Markowitz, K. Melton, B. Morey, C. Otoul, S. Patel, J. Phillips, E. Sen-Gupta, O. Stumpp, D. Tatla, D. Terricabras, K. Claes, J.A. Wright, N. Sheth, Clinical feasibility of a wearable, conformable sensor patch to monitor motor symptoms in Parkinson’s disease, Park. Relat. Disord. 61 (2019) 70–76. https://doi.org/10.1016/j.parkreldis.2018.11.024.

[27] D. Berg, R.B. Postuma, C.H. Adler, B.R. Bloem, P. Chan, B. Dubois, T. Gasser, C.G. Goetz, G. Halliday, L. Joseph, A.E. Lang, I. Liepelt-Scarfone, I. Litvan, K. Marek, J. Obeso, W. Oertel, C.W. Olanow, W. Poewe, M. Stern, G. Deuschl, MDS research criteria for prodromal Parkinson’s disease, Mov. Disord. 30 (2015) 1600–1611. https://doi.org/10.1002/mds.26431.

[28] A. Mirelman, T. Gurevich, N. Giladi, A. Bar-Shira, A. Orr-Urtreger, J.M. Hausdorff, Gait alterations in healthy carriers of the LRRK2 G2019S mutation, Ann. Neurol. (2011). https://doi.org/10.1002/ana.22165.

[29] E.M. Mcdade, B.P. Boot, T.J.H. Christianson, V.S. Pankratz, B.F. Boeve, T.J. Ferman, K. Bieniek, J.H. Hollman, R.O. Roberts, M.M. Mielke, D.S. Knopman, R.C. Petersen, Subtle gait changes in patients with REM sleep behavior disorder, Mov. Disord. 28 (2013) 1847–1853. https://doi.org/10.1002/mds.25653.

[30] S. Del Din, M. Elshehabi, B. Galna, M.A. Hobert, E. Warmerdam, U. Suenkel, K. Brockmann, F. Metzger, C. Hansen, D. Berg, L. Rochester, W. Maetzler, Gait analysis with wearables predicts conversion to parkinson disease, Ann. Neurol. 86 (2019) 357–367. https://doi.org/10.1002/ana.25548.

[31] A.J. Espay, L. V. Kalia, Z. Gan-Or, C.H. Williams-Gray, P.L. Bedard, S.M. Rowe, F. Morgante, A. Fasano, B. Stecher, M.A. Kauffman, M.J. Farrer, C.S. Coffey, M.A. Schwarzschild, T. Sherer, R.B. Postuma, A.P. Strafella, A.B. Singleton, R.A. Barker, K. Kieburtz, C.W. Olanow, A. Lozano, J.H. Kordower, J.M. Cedarbaum, P. Brundin, D.G. Standaert, A.E. Lang, Disease modification and biomarker development in Parkinson disease: Revision or reconstruction?, Neurology. 94 (2020) 481–494. https://doi.org/10.1212/WNL.0000000000009107.

[32] A.J. Espay, P. Bonato, F.B. Nahab, W. Maetzler, J.M. Dean, J. Klucken, B.M. Eskofier, A. Merola, F. Horak, A.E. Lang, R. Reilmann, J. Giuffrida, A. Nieuwboer, M. Horne, M.A. Little, I. Litvan, T. Simuni, E.R. Dorsey, M.A. Burack, K. Kubota, A. Kamondi, C. Godinho, J.F. Daneault, G. Mitsi, L. Krinke, J.M. Hausdorff, B.R. Bloem, S. Papapetropoulos, Technology in Parkinson’s disease: Challenges and opportunities, Mov. Disord. (2016). https://doi.org/10.1002/mds.26642.

